# Poor sleep quality, insomnia, and short sleep duration before infection predict long-term symptoms after COVID-19

**DOI:** 10.1101/2023.02.13.23285859

**Authors:** Federico Salfi, Giulia Amicucci, Domenico Corigliano, Lorenzo Viselli, Aurora D’Atri, Daniela Tempesta, Michele Ferrara

## Abstract

**Study objectives:** Millions of COVID-19 survivors experience a wide range of long-term symptoms after acute infection, giving rise to serious public health concerns. To date, few risk factors for post-COVID-19 conditions have been determined. This study evaluated the role of pre-infection sleep quality/duration and insomnia severity in the incidence of long-term symptoms after COVID-19.

**Methods:** This prospective study involved two assessments (April 2020 and 2022). At baseline, sleep quality/duration and insomnia symptoms in participants without current/prior SARS-CoV-2 infection were measured using the Pittsburgh Sleep Quality Index (PSQI) and the Insomnia Severity Index (ISI). At follow-up, we evaluated the presence of twenty-one symptoms (psychiatric, neurological, cognitive, bodily, and respiratory) one month (n=713, infection in April 2020–February 2022) and three months after COVID-19 (n=333, infection in April 2020–December 2021). Zero-inflated negative binomial models were used to estimate the effect of previous sleep on the number of long-term symptoms. Binomial logistic regressions were performed to evaluate the association between sleep outcomes and the incidence of each post-COVID-19 symptom.

**Results:** Analyses highlighted a significant effect of pre-infection sleep on the number of symptoms one/three months after COVID-19. Previous higher PSQI and ISI scores, and shorter sleep duration significantly increased the risk of almost every long-term symptom at one/three months from COVID-19.

**Conclusion:** This study suggested a prospective dose-dependent association between pre-infection sleep quality/quantity and insomnia severity with the manifestation of post-COVID-19 symptoms. Promoting sleep health may represent an effective preventive approach to mitigate the COVID-19 sequelae, with substantial public health and societal implications.

**Statement of significance:** Determining potential risk factors of long COVID is crucial to driving preventive interventions amongst vulnerable populations. The present study is the first to provide insights into the role of pre-infection sleep disturbances in the occurrence of post-COVID-19 conditions. We demonstrated a strong association between previous sleep quality/duration and insomnia severity with the incidence of a broad spectrum of long-term symptoms one and three months after COVID-19. These results may have large-scale implications, considering the alarming rates of both sleep disturbances and post-COVID-19 manifestations worldwide. Further research is warranted to determine the biobehavioral mechanisms involved. Future studies should also evaluate whether treating sleep problems may improve the subsequent long-term consequences of COVID-19.

## Introduction

Estimates from the World Health Organization indicate that over 750 million Coronavirus Disease 2019 (COVID-19) cases have been confirmed globally.^1^ While most individuals experience mild symptoms and recover quickly, recent meta-analyses suggested that 43–45% of COVID-19 survivors report signs and symptoms that continue or develop in the long run.^2,3^ This condition is commonly termed “long COVID” and encompasses over two-hundred long-term clinical manifestations.^4^ General symptoms include fatigue, body aches, fever, ageusia, and anosmia. Other symptoms are related to lung disease (i.e., cough, dyspnea) or involve neurological and cognitive dysfunctions (headache, brain fog, attention/concentration disorders, memory loss) and cardiovascular or gastrointestinal disorders. Moreover, a broad spectrum of psychiatric and psychological manifestations after acute infection were also identified, such as sleep disorders, depression, anxiety, post-traumatic stress disorder (PTSD), obsessive-compulsive disorders (OCD) and psychosis.^5^ This disabling condition pervasively impacts the daily life of COVID-19 survivors, also affecting their ability to resume a regular working routine.^6,7^

Identifying potential antecedents of long-term symptoms represents a first-order medical challenge due to the burden on the international healthcare systems and the societal and economic costs.^8^ However, due to the multisystemic and heterogeneous nature of long COVID, its etiology remains poorly understood, and current evidence propose chronic inflammation and immune dysregulation as possible causes of long-term clinical manifestations after acute illness.^9–12^

Sleep plays a crucial role in human immunity,^13,14^ and poor sleep quality and inadequate sleep duration are associated with increased susceptibility to virus infections.^15,16^ Moreover, a growing body of evidence linked sleep disturbances and short sleep duration with increased risk for inflammatory diseases due to the relationship between sleep problems and low sleep amount with sustained production of pro-inflammatory cytokines and other circulating markers of inflammation.^17,18^

Based on these assumptions, pre-infection sleep disturbances could play a role in predisposing people to experience long-term symptoms after COVID-19. However, to the best of our knowledge, no prospective studies have addressed this research question. In this study, sleep outcomes of validated questionnaires from a nationwide survey held during the first Italian lockdown (April 2020) were used as predictors of long COVID symptoms in a group who reported a positive swab for SARS-CoV-2 in the subsequent two years. We hypothesized that sleep disturbances and shorter sleep duration could be prospectively associated with the occurrence of a wide range of long-term symptoms after one and three months from COVID-19 while accounting for established risk factors (age, gender, body mass index – BMI, COVID-19 severity).^2,19–21^ Finally, we investigated whether sleep issues before infection were related to longer recovery times to return to the pre-infection daily functioning level.

## Materials and methods

### Participants and procedure

A total of 13,989 participants were surveyed during the first lockdown period in April 2020 via a web-based set of questionnaires (for a detailed description of the data collection procedure, see^22^). Subsequently, a total of 2,013 respondents were longitudinally evaluated in December 2020.^23^ Finally, the overall sample surveyed in April 2020 was re-invited to take part in another longitudinal assessment in April 2022. A total of 2,759 Italians participated in the last data collection, while a total of 1,062 respondents participated in all three survey waves.^24^ Each assessment comprised an evaluation of sleep quality/duration and insomnia severity using the Pittsburgh Sleep Quality Index^25^ (PSQI), and the Insomnia Severity Index^26^ (ISI).

The PSQI is a reliable questionnaire to evaluate sleep quality.^25^ It consists of nineteen questions, from which a total score (range, 0–21) is calculated. A higher score indicates poorer sleep quality. The ISI is a validated screening instrument to assess the severity of insomnia symptoms.^26^ It comprised seven items and a higher total score (range, 0–28) point to more severe insomnia.

Moreover, in each survey wave we collected demographic and other information (for more details, see^22–24^). In April 2022, we asked respondents if they have ever tested positive for COVID-19. If so, participants were asked to answer a set of *ad hoc* questions about their infection and symptomatology. We collected information about:

I. the month of detected swab positivity;
II. the COVID-19 severity in the acute stage of illness (*no marked symptoms*: absence of any symptom except for smell/taste dysfunctions; *mild disease*: e.g., cough, fever, muscle pains, etc., without pneumonia; *moderate disease*: non-severe pneumonia and having received different medications without the need of extra oxygen treatment; *severe disease*: severe pneumonia requiring extra-oxygen therapy and intravenous lines attached);
III. the recovery time (in weeks) to return to the pre-infection daily functioning level;
IV. the presence of long-term symptoms one and three months after the first infection according to Italian National Institute of Health guidelines for long-COVID identification (over-tiredness, muscle weakness, breathlessness/dyspnea, concentration/attention difficulty, headache, asthenia, anxiety, diffuse body pain, sleep problems, memory problems, brain fog, deterioration of perceived health status, persistent cough, smell/taste dysfunctions, depression, appetite reduction, fever, PTSD, cardiovascular problems, OCD, psychosis).

From the overall April 2022 sample, a total of 973 participants (35.00%, mean age ± standard deviation, 33.40 ± 11.40 years; range, 18–81 years; 170 males) reported SARS-CoV-2 infection and provided the above-listed information. The temporal distributions of COVID-19 cases across the pandemic period in our sample and among the over 18-year Italian population are reported in Figure 1.

**Figure 1.**
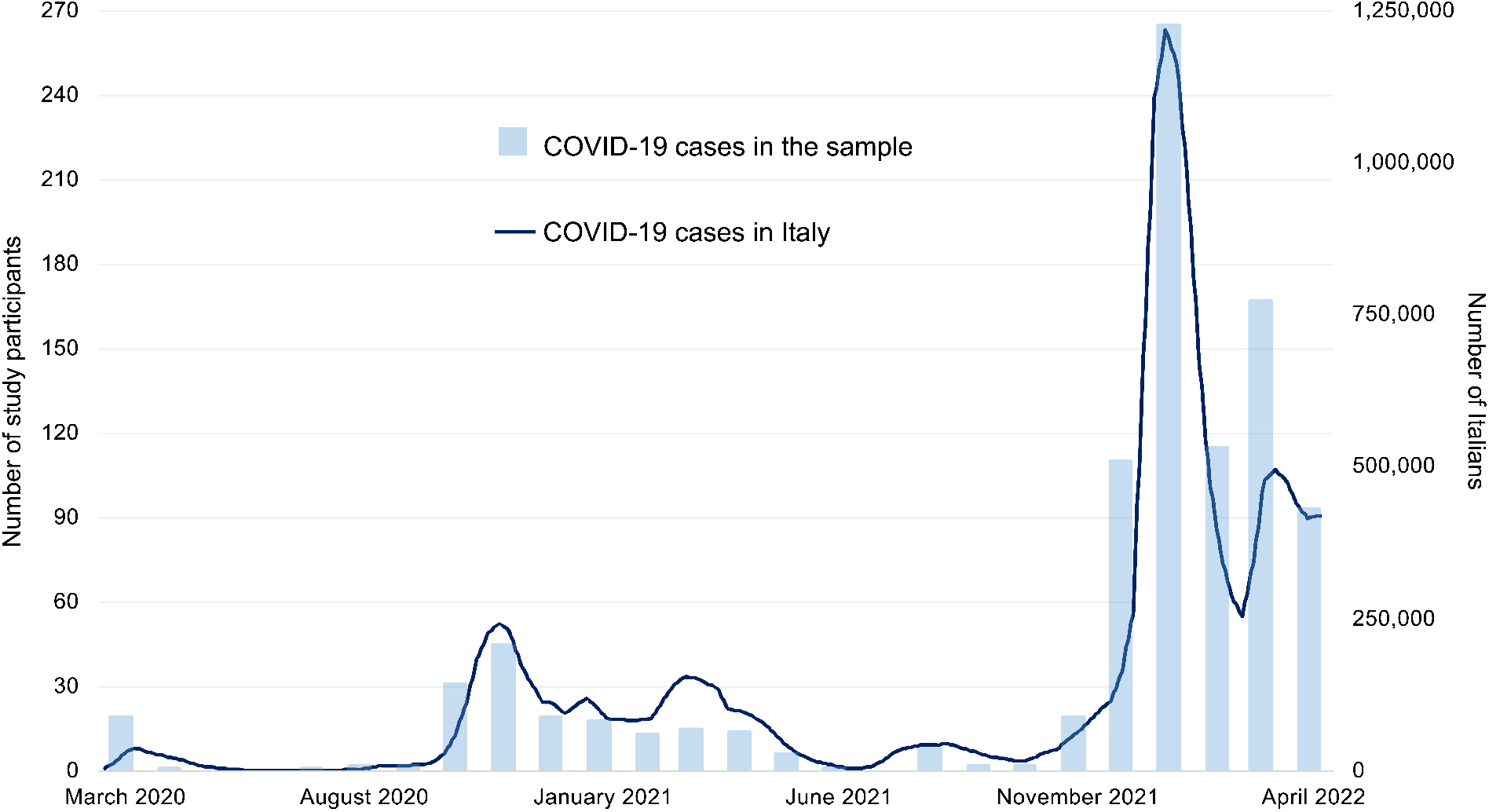
Temporal distribution of COVID-19 cases among the over 18-year Italian population (weekly; dark blue line) and in the overall study sample (n= 973; monthly; light blue bars).

To estimate the role of sleep quality, insomnia severity, and total sleep time (TST, extracted from the item 4 of PSQI) in predicting long-term symptoms one month from COVID-19, the analyses were performed on 713 people (33.38 ± 11.40 years; 18–81 years; 122 males) who reported the positivity from April 2020 to February 2022, excluding subjects whose infection was less than a month old during the third survey wave or people already infected during the first survey wave. To evaluate the effect of sleep variables in predicting long-term symptoms at three months from the infection, the analyses were performed on 333 individuals (33.09 ± 11.80 years; 18–70 years; 60 males) who reported a swab positivity from February 2020 to December 2021, excluding subjects whose infection was less than three months old in April 2022 or people reporting infection before/during the first assessment (April 2020). The flow chart of participants analyzed in the study is reported in Figure S1 in the supplemental material.

The project was approved by the Institutional Review Board of the University of L’Aquila (protocol no. 43066). Respondents provided electronic consent to participate in the study in each survey wave.

## Statistical analysis

We planned three analysis families to evaluate the predictive effect of the sleep variables on (i) the number of long-term symptoms, (ii) the odds of each long-term symptom, and (iii) the odds of recovering the pre-infection daily functioning level in longer times (after 4 or 12 weeks). All analyses described in the following sections were adjusted for demographic confounding variables [age, gender, and BMI (weight/height^2^)]. All tests were two-tailed and a *p*-value < 0.05 was considered significant.

A set of control analyses were also performed to evaluate the consistency of our results. Firstly, all analyses were further adjusted for the COVID-19 severity, considering the established relationship between acute illness severity and future long-covid symptoms.^2,19,20,27^ Secondly, models were fitted including the number of months between April 2020 and the time of the infection as covariate to control for the time distance between actual infection and the first survey wave. Finally, analyses were performed also using a different baseline assessment of sleep variables (December 2020; see “Supplemental statistical analysis” section in the supplemental material).

### Analysis on the number of long-term symptoms after COVID-19

The numbers of symptoms at one/three months from COVID-19 were entered as dependent variables (range, 0–21), and the sleep variables (PSQI score, ISI score, TST) as separate predictors. Due to the nature of the dependent variables (count data), linear regressions are unsuitable.^28,29^ Therefore, we evaluated the possibility of performing Poisson regressions, which are typically used to model count data.^28,29^ However, preliminary analyses highlighted a violation of the equidispersion assumption^29^ (*χ*^2^/*df* ranging from 3.94 to 5.48, greater than the acceptable value of 1.20^30^). In this case, a Negative Binomial (NB) regression approach should be used.^28,31,32^ NB models provide estimates [*exp(B)*] that indicate the rate change of the count variable for a one-unit change in the predictor.

Due to the high prevalence of zero values in count variables (27.9% and 52.6% in one-month and three-month symptoms, respectively), data were modeled with Zero-Inflated NB regression^33–35^ (ZINB). The ZINB approach combines two separate models: a ZI model and a NB model. The ZI portion is used to model the probability of a zero count, while the negative binomial portion is used to model the count itself. The ZINB model is useful in analyzing count data that has an excess of zero values because it accounts for this excess in the data and provides more accurate estimates of the underlying distribution. Therefore, the ZINB models allowed us to manage both the excess of zero observations and the overdispersion in the data.

Six ZINB models (M) were tested, evaluating the effect of PSQI, ISI, and TST in predicting the number of symptoms at one month (M1, M2, M3, respectively) and three months from infection (M4, M5, M6, respectively). The models were fitted using “*countereg*” R package.^36^ In the results section, we focused on the NB portion of ZINB models for addressing the role of sleep variables in predicting the number of long-term symptoms. A comparison of relative goodness-of-fit (Akaike information criterion and the Bayesian information criterion) supported the use of ZINB regressions instead of NB (see Table S1 in the supplemental material). Moreover, a visual inspection of Tukey’s rootograms^37^ using the “*countereg*” R package^38^ confirmed ZINB as the best-fit models.

### Analysis on each long-term symptom after COVID-19

Binomial logistic regressions were performed to evaluate the association between sleep variables and the future occurrence of each long-term symptom one and three months after COVID-19. Analyses were performed using “*glm*” function of “*stats*” R package.^39^ In detail, each sleep variable was entered as a predictor (PSQI score, ISI score, TST), and each long-term symptom as a dichotomous dependent variable (yes, no). The criterion of a minimum of 5 outcome events per predictor variable (EPV) was adopted^40^ to identify the analyzable long-term symptoms after infection. Consequently, some symptoms were excluded from the analysis on one-month (psychosis and OCD) and three-month symptoms (PTSD, fever, persistent cough, OCD, appetite reduction, cardiovascular problems, and psychosis) due to insufficient events. Considering the relaxed EPV criterion adopted to maximize the number of analyzable long-term symptoms, sensitivity analyses were performed replicating the binomial logistic models using the Firth’s bias reduction method.^41^ This approach allows the parameter estimations to be more efficient and robust to small sample sizes and rare events by penalizing the likelihood function. These control analyses were applied using “*logistf”* R package.^42^

### Analysis on recovery time after COVID-19

Binomial logistic regressions were performed to test the association between sleep characteristics (quality, duration, insomnia severity) and the recovery time to return to the pre-infection daily functioning level. The self-reported recovery time was entered as dichotomous dependent variable (≤ 4 weeks vs. > 4 weeks for the one-month symptoms sample; ≤ 12 weeks vs. > 12 weeks for the three-month symptoms sample), while sleep variables (PSQI score, ISI score, TST) were used as predictors.

## Results

### Sample characteristics and COVID-19-related information

The composition of the analyzed samples is reported in Table 1. Most participants were young, females, fell within the healthy weight range, and had at least a bachelor’s degree. The majority of respondents reported a mild COVID-19 severity in the acute stage and recovered the pre-infection daily functioning level in less than a month. Descriptive statistics of questionnaire scores assessing sleep quality, severity of insomnia symptoms, and sleep duration are also reported.

**Table 1.** Demographic characteristics, information about COVID-19, and descriptive statistics of questionnaire scores evaluating sleep quality, insomnia severity, and sleep duration among participants who were tested positive for SARS-CoV-2 from April 2020 to February 2022, and from April to December 2021.

| Variable | COVID-19 in<br>April 2020–February 2022<br>(n = 713) | COVID-19 in<br>April 2020–December 2021<br>(n = 333) |
| --- | --- | --- |
|  | N (%) or *mean (SD) |  |
| Age |  |  |
| Younger (18 to 30 years) | 390 (54.7) | 194 (58.3) |
| Middle-aged (31 to 50 years) | 250 (35.1) | 100 (30.0) |
| Older (> 50 years) | 73 (10.2) | 39 (11.7) |
| Gender |  |  |
| Male | 122 (17.1) | 60 (18.0) |
| Female | 591 (82.9) | 273 (82.0) |
| BMI |  |  |
| Underweight range (< 18.5) | 48 (6.7) | 23 (6.9) |
| Healthy weight range (18.5 to < 25) | 462 (64.8) | 205 (61.6) |
| Overweight range (25 to < 30) | 146 (20.5) | 69 (20.7) |
| Obesity (≥ 35) | 57 (8.0) | 36 (10.8) |
| Education |  |  |
| Middle/High school | 216 (30.3) | 106 (31.8) |
| Bachelor's degree | 171 (24.0) | 108 (32.4) |
| Master's degree | 229 (32.1) | 78 (23.4) |
| Postgraduate | 97 (13.6) | 41 (12.3) |
| COVID-19 severity |  |  |
| No marked symptoms | 183 (25.7) | 84 (25.2) |
| Mild disease | 490 (68.7) | 220 (66.1) |
| Moderate disease | 33 (4.6) | 22 (6.6) |
| Severe disease | 7 (1.0) | 7 (2.1) |
| Time to recover the pre-infection daily functioning level |  |  |
| ≤ 4 weeks | 479 (67.2) |  |
| > 4 weeks | 234 (32.8) |  |
| ≤ 12 weeks |  | 270 (81.1) |
| > 12 weeks |  | 63 (18.9) |
| PSQI score | *6.950 ± 3.575 | *6.885 ± 3.694 |
| ISI score | *8.279 ± 5.419 | *8.171 ± 5.330 |
| Sleep duration (hour) | *7.205 ± 1.358 | *7.215 ± 1.422 |

Notes: Numbers preceded by an asterisk (\*) are mean values (standard deviation). BMI ranges were established according to the American Centers for Disease Control and Prevention guidelines.<sup>43</sup>
Abbreviations: BMI, Body mass index; ISI, Insomnia Severity Index; PSQI, Pittsburgh Sleep Quality Index; SD, Standard deviation.

The prevalence of each long-term symptom one month and three months after COVID-19 among the analyzed samples is reported in Figure 2a. The distribution of the number of symptoms at one month and three months from the infection is shown in Figure 2b.

**Figure 2.**
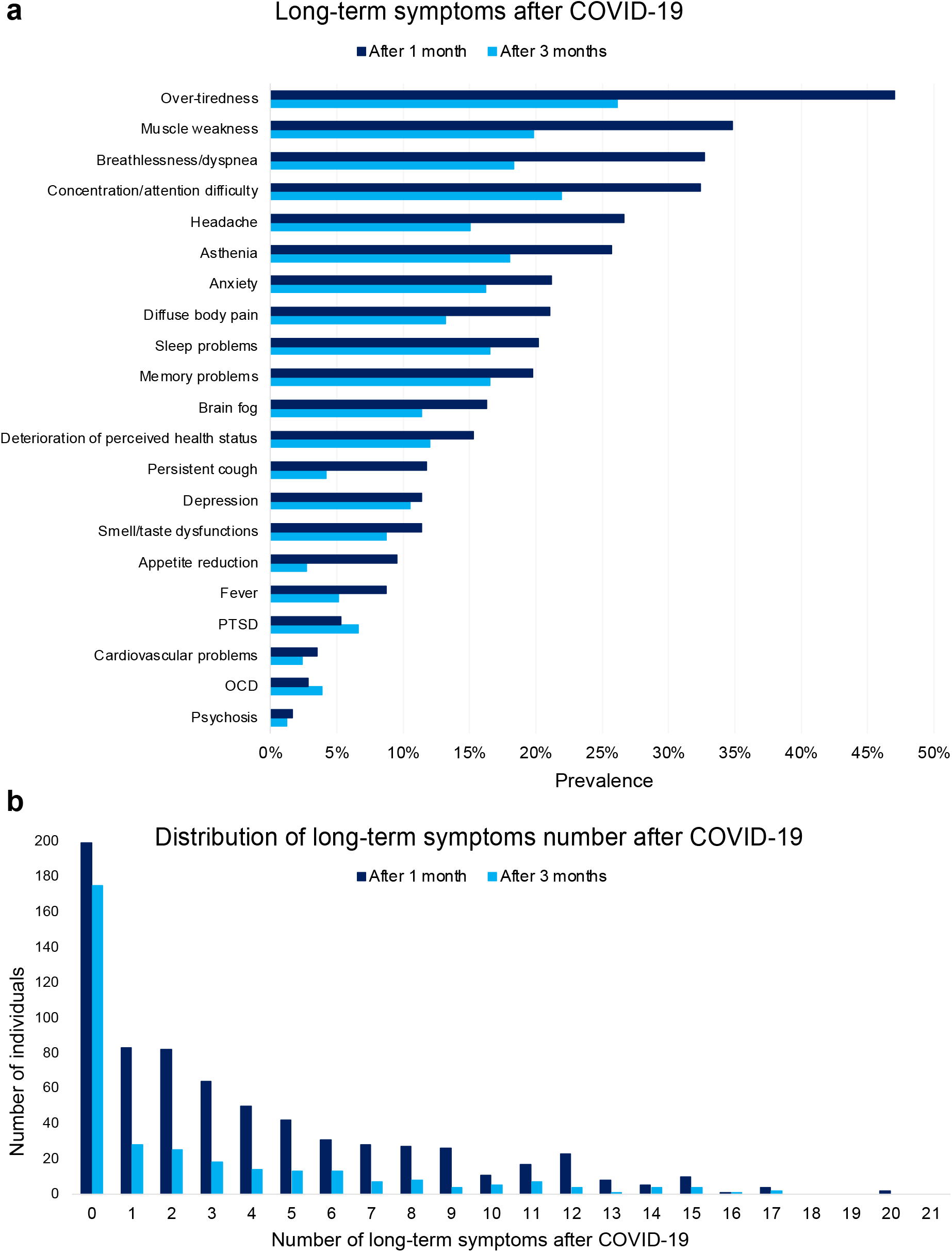
Prevalence of long-term symptoms (a) and distribution of symptoms number (b) one month and three months after COVID-19 in the analyzed samples. *Notes:* The “After 1 month” group consists of 713 individuals (dark blue bars), while the “After 3 months” group comprises 333 subjects (light blue bars). *Abbreviations:* OCD, obsessive-compulsive disorder; PTSD, post-traumatic stress disorder.

### Association between pre-infection sleep and the number of long-term symptoms after COVID-19

Results from the NB portion of the ZINB models (Table 2) showed that lower sleep quality, more severe insomnia symptoms, and shorter sleep duration significantly predicted a higher number of long-term symptoms at one month and three months after COVID-19. A one-unit increase in PSQI and ISI scores, and a one-hour reduction of sleep duration predicted a 7.0%, 4.9%, and 11.1% increase in the number of symptoms one month after infection, and an increase of 9.1%, 5.4%, and 14.7% in the number of symptoms three months after COVID-19, respectively. A graphical representation of the relationships between sleep variables and the number of long-term symptoms is reported in Figure 3.

**Table 2.** Results from the negative binomial portion of the zero-inflated negative binomial regressions [*exp(B)*, 95% confidence intervals, *p*-value] estimating the effect of sleep variables (PSQI score, ISI score, TST) and confounding factors (age, gender, BMI) in April 2020 on the number of long-term symptoms one month (M1, M2, M3) and three months after COVID-19 (M4, M5, M6).

| Predictor | M1: sleep quality |  |  | M2: insomnia severity |  |  | M3: sleep duration |  |  |
| --- | --- | --- | --- | --- | --- | --- | --- | --- | --- |
| | $\exp(B)$ | 95% CI | $p$ | $\exp(B)$ | 95% CI | $p$ | $\exp(B)$ | 95% CI | $p$ |
| Intercept | 2.325 | 1.461–3.699 | **< 0.001** | 2.315 | 1.465–3.658 | **< 0.001** | 9.182 | 4.494–18.761 | **< 0.001** |
| Gender* | 0.912 | 0.724–1.149 | 0.433 | 0.878 | 0.701–1.101 | 0.260 | 0.819 | 0.649–1.034 | 0.093 |
| Age | 0.990 | 0.983–0.998 | **0.009** | 0.992 | 0.984–0.999 | **0.030** | 0.987 | 0.979–0.995 | **0.001** |
| BMI | 1.023 | 1.006–1.041 | **0.008** | 1.024 | 1.006–1.041 | **0.008** | 1.026 | 1.008–1.045 | **0.004** |
| PSQI score | 1.070 | 1.046–1.094 | **< 0.001** |  |  |  |  |  |  |
| ISI score |  |  |  | 1.049 | 1.033–1.064 | **< 0.001** |  |  |  |
| TST (hour) |  |  |  |  |  |  | 0.889 | 0.840–0.941 | **< 0.001** |
| Predictor | M4: sleep quality |  |  | M5: insomnia severity |  |  | M6: sleep duration |  |  |
| | $\exp(B)$ | 95% CI | $p$ | $\exp(B)$ | 95% CI | $p$ | $\exp(B)$ | 95% CI | $p$ |
| Intercept | 1.312 | 0.551–3.124 | 0.540 | 1.706 | 0.776–3.752 | 0.184 | 8.668 | 2.478–30.317 | **0.001** |
| Gender* | 1.021 | 0.592–1.446 | 0.733 | 1.017 | 0.644–1.550 | 0.997 | 1.027 | 0.522–1.278 | 0.375 |
| Age | 1.002 | 0.987–1.018 | 0.772 | 1.002 | 0.988–1.017 | 0.746 | 0.996 | 0.980–1.012 | 0.628 |
| BMI | 0.925 | 0.990–1.054 | 0.184 | 0.999 | 0.986–1.048 | 0.284 | 0.816 | 0.995–1.059 | 0.097 |
| PSQI score | 1.091 | 1.047–1.136 | **< 0.001** |  |  |  |  |  |  |
| ISI score |  |  |  | 1.054 | 1.027–1.082 | **< 0.001** |  |  |  |
| TST (hour) |  |  |  |  |  |  | 0.853 | 0.769–0.946 | **0.003** |

Notes: \*Female was used as reference for “Gender” factor; significant values are in bold.
Abbreviations: BMI, Body mass index; CI, Confidence interval; ISI, Insomnia Severity Index; M, Model; PSQI, Pittsburgh Sleep Quality Index; TST, Total sleep time.

**Figure 3.**
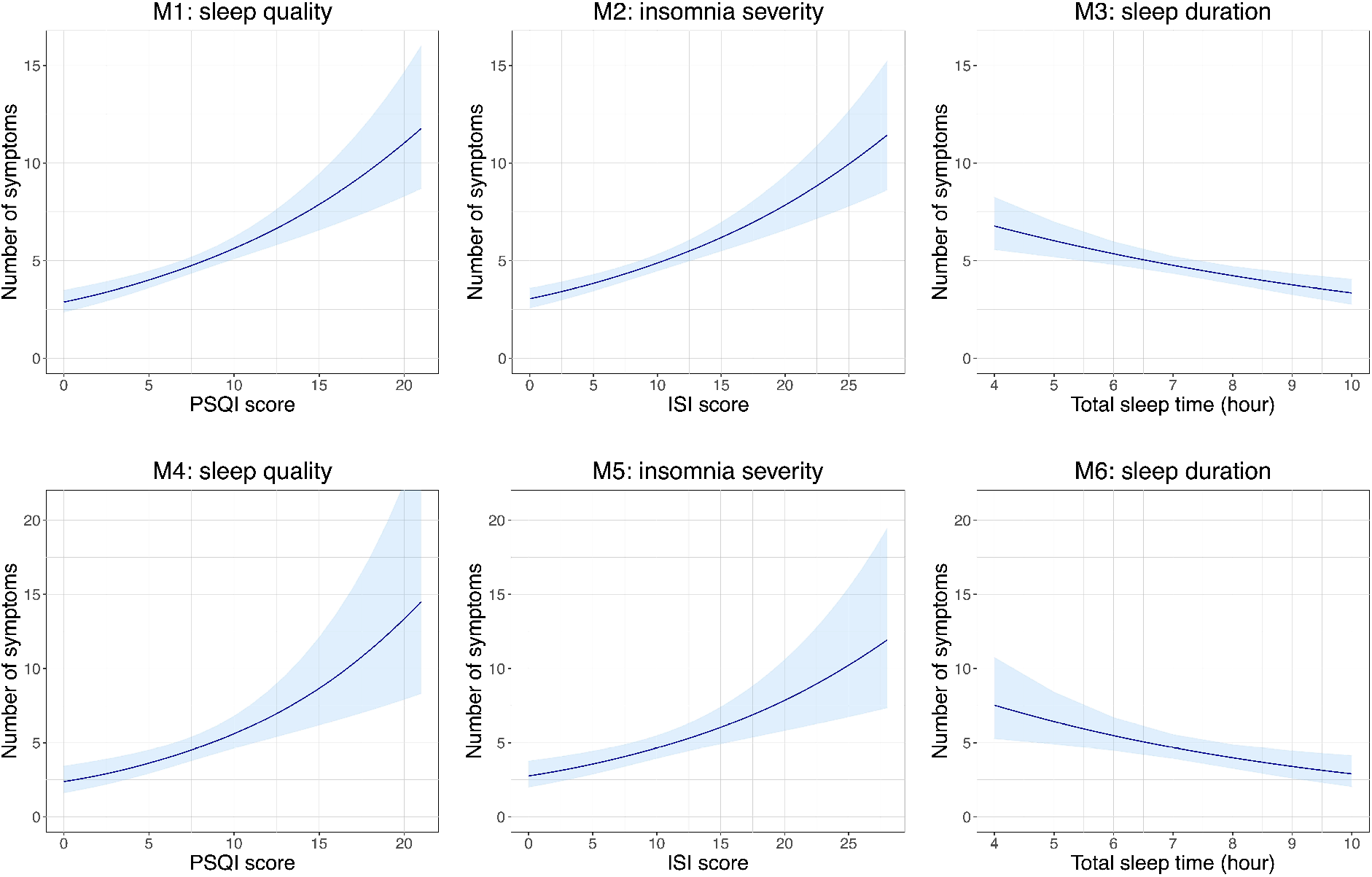
Relationships between sleep variables (PSQI score, ISI score, TST) in April 2020 and the number of long-term symptoms one month (M1, M2, M3) and three months after COVID-19 (M4, M5, M6). *Notes:* Light blue area represents 95% confidence intervals. Each model was adjusted for age, gender, and body mass index. *Abbreviations:* ISI, Insomnia Severity Index; M, Model; PSQI, Pittsburgh Sleep Quality Index; TST, Total sleep time.

Sensitivity analyses adjusting M1–6 for the COVID-19 severity or the time distance between swab positivity and the baseline assessment (April 2020) confirmed all the significant effects of sleep variables. Results were also confirmed using sleep outcomes collected in December 2020 as predictors of one-month symptoms (see Table S2 and Figure S2 in the supplemental material).

Finally, supplementary analyses showed that the occurred variations in sleep quality, insomnia severity, and sleep duration between April and December 2020 significantly predicted the number of long-term symptoms one month after infection (see Table S3 and Figure S3 in the supplemental material).

### Association between pre-infection sleep and each long-term symptom after COVID-19

Results of binomial logistic regressions using sleep variables collected in April 2020 as predictors indicated that higher PSQI scores significantly increased the odds of each analyzed long-term symptom at one (Figure 4a) and three months from COVID-19 (Figure 4b), except for smell/taste dysfunctions, and cardiovascular problems experienced at one month from infection. More severe insomnia symptoms were significantly associated with higher odds of all analyzed long-term symptoms, excluding smell/taste dysfunctions Figure 4c,d). Finally, a one-hour decrease in sleep duration was associated with higher odds of all one-month long-term symptoms, except for asthenia, memory problems, smell/taste dysfunctions, appetite reduction, and cardiovascular problems (Figure 4e). Furthermore, reduced sleep duration was associated with increased risk for all symptoms reported after 3 months from infection, excluding over-tiredness, concentration/attention difficulty, anxiety, depression, and smell/taste dysfunctions (Figure 4f).

**Figure 4.**
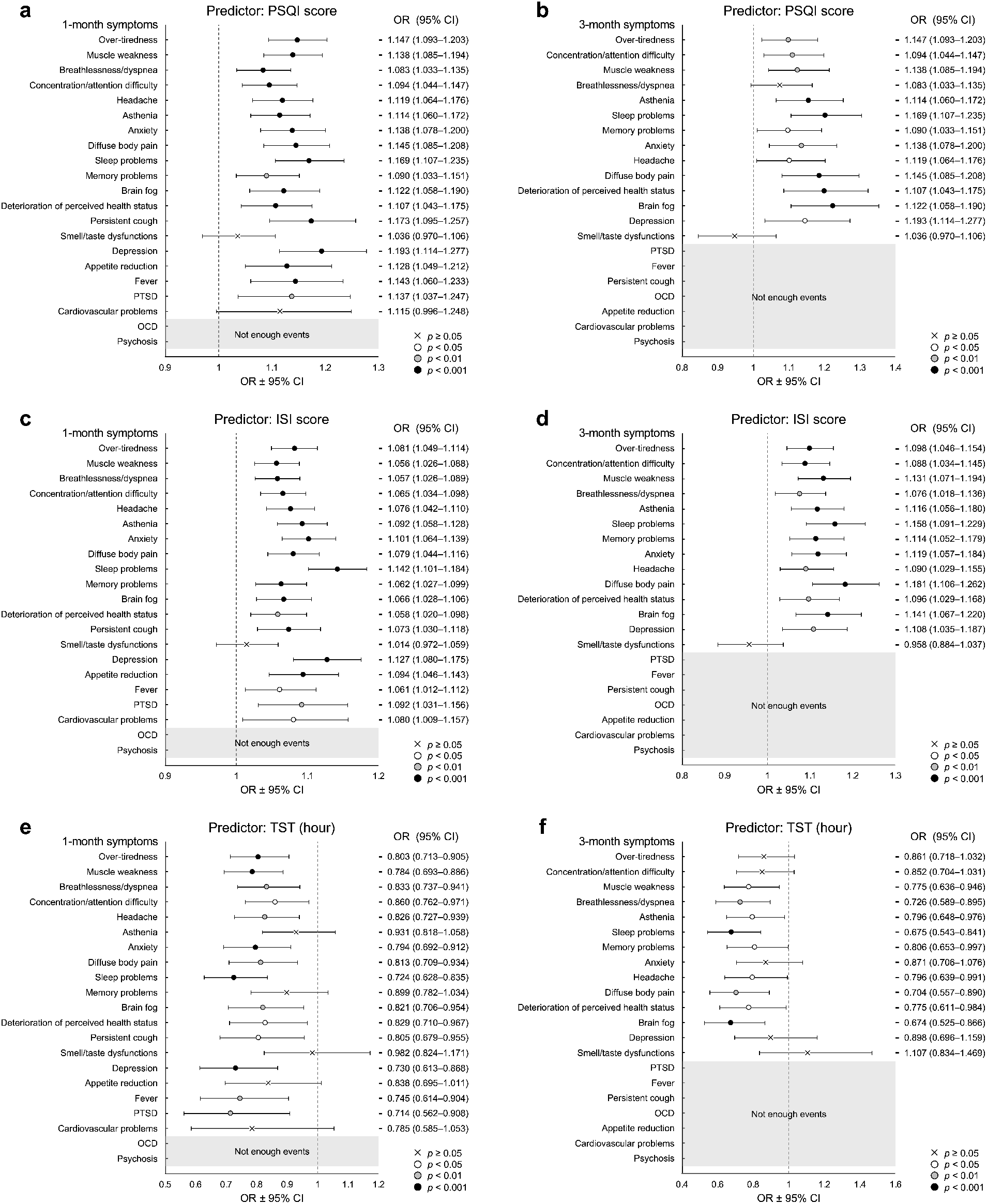
Results of logistic regressions (odd ratios and 95% confidence intervals) evaluating the predictive effect of sleep variables (PSQI score, ISI score, TST) in April 2020 on the odds of each long-term symptom one month (a, c, e) and three months after COVID-19 (b, d, f). *Notes:* Long-term symptoms were ordered according to the prevalence data (top: most frequent) and represent the dependent variables. White dot indicates significance level at *p* < 0.05, grey dot at *p* < 0.01, and black dot at *p* < 0.001. “×” symbol indicates no statistically significant effect. Grey area indicates insufficient (< 5) outcome events per predictor. Each model was adjusted for age, gender, and body mass index. *Abbreviations:* CI, Confidence Interval; ISI, Insomnia Severity Index; OCD, obsessive-compulsive disorder; OR, odd ratio; PSQI, Pittsburgh Sleep Quality Index; PTSD, post-traumatic stress disorder; TST, Total sleep time.

Control analyses using Firth’s bias-reduced logistic regressions produced almost identical results to those obtained using the reported binomial logistic regressions, rejecting possible bias due to the low number of events per predictor for some long-term symptoms. Similarly, adjusting for self-reported COVID-19 severity or the time distance between SARS-CoV-2 infection and the baseline assessment (April 2020) confirmed the overall pattern of results.

Finally, logistic models using PSQI score, ISI score, and TST of December 2020 as predictors of symptoms reported one month after COVID-19 partially confirmed the above-described results (see Figure S4 in the supplemental material).

### Association between pre-infection sleep and recovery time after COVID-19

Binomial logistic regression showed that poor sleep quality, more severe insomnia symptoms, and shorter sleep duration in April 2020 predicted longer recovery time to return to the pre-infection daily functioning level after COVID-19 (Figure 5). A one-unit increase in PSQI and ISI scores, and a one-hour reduction of sleep duration were prospectively associated with higher odds of recovery in more than four weeks by 13.1%, 9.3%, and 14.4%, and after twelve weeks by 21.3%, 12.0%, and 20.2%, respectively. Adjusting for COVID-19 severity and the time distance between April 2020 assessment and the virus infection confirmed the above results. Finally, logistic regressions using sleep variables collected in December 2020 as predictors confirmed the significant association between recovery after 4 weeks with PSQI and ISI scores (see supplemental material).

**Figure 5.**
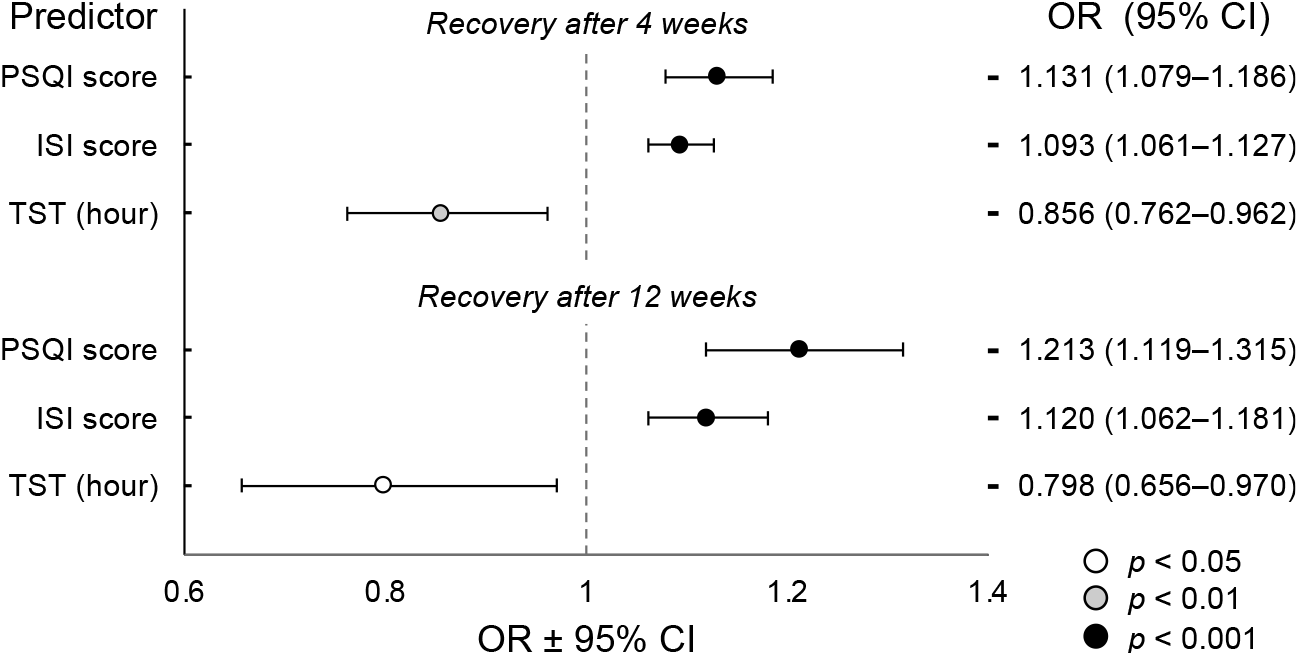
Results of logistic regressions (odd ratios and 95% confidence intervals) evaluating the predictive effect of sleep variables (PSQI score, ISI score, TST) in April 2020 on the odds of returning to the pre-infection daily functioning level after 4 and 12 weeks. *Notes:* Self-reported recovery after 4 and 12 weeks was evaluated on 713 and 333 subjects, respectively. White dot indicates significance level at *p* < 0.05, grey dot at *p* < 0.01, and black dot at *p* < 0.001. Each model was adjusted for age, gender, and body mass index. *Abbreviations:* CI, Confidence Interval; ISI, Insomnia Severity Index; OR, Odd ratio; PSQI, Pittsburgh Sleep Quality Index; TST, Total sleep time.

## Discussion

Since the beginning of the pandemic, millions of people worldwide have reported signs and symptoms that continue or develop after COVID-19, affecting their ability to resume normal life and giving rise to serious public health concerns.^8^

To the best of our knowledge, this study is the first to find a prospective association between pre-infection sleep disturbances and the occurrence of long-term symptoms after COVID-19. We highlighted significant dose-dependent relationships between previous sleep quality, insomnia severity, and sleep duration and the number of symptoms experienced at one and three months from the reported positive swab for SARS-CoV-2. Moreover, our study showed that lower sleep quality, more severe insomnia, and shorter sleep time lead to higher odds of experiencing a broad spectrum of clinical manifestations one and three months after COVID-19. Finally, we found that previous sleep problems are related to longer recovery times for returning to the self-perceived pre-infection daily functioning.

Our results were consistent with a recent study on a sample of female nurses that found an association between short sleep duration collected in 2017 and the occurrence of post-COVID-19 conditions.^44^

Immune dysregulation and inflammatory mechanisms may explain the association between sleep and subsequent post-COVID-19 manifestations. Consistent literature showed that sleep disturbances are associated with heightened systemic inflammation, as evidenced by increased levels of pro-inflammatory cytokines,^17,45–48^ C-reactive protein,^17,47–50^ and other markers of inflammation.^46,51^ This evidence has been proposed to explain the documented higher risk for inflammatory diseases in people with sleep disturbances.^17,18^ On the other hand, findings from animal^52,53^ and human models^9–12^ suggested that a possible mechanism behind the long COVID symptoms may involve an abnormal and persistent pro-inflammatory response weeks/months after infection. For example, Phetsouphanh and collaborators^9^ compared immune profiles of long COVID individuals with patients without long COVID, identifying a combination of inflammatory mediators eight months after infection as the correlate of persistent post-COVID-19 symptoms.

Another interpretation may involve the role of sleep loss as a driver of cellular stress and the consequent neuronal damage^54^ in the cognitive and neuropsychiatric manifestations of long COVID.^55^

Finally, pre-pandemic evidence suggested that insufficient sleep could impair the vaccination efficacy,^56–58^ and the importance of sleep health has also been advocated for the COVID-19 vaccination campaign.^59,60^ Since previous COVID-19 vaccination seems to reduce the risk of long COVID,^61^ the link between sleep and post-COVID-19 symptoms may be mediated by the impact of sleep deficiency on the vaccination effectiveness. However, providing a clear understanding of the link between sleep and subsequent long-term symptoms after SARS-CoV-2 infection transcends the objectives and the capacity of the present investigation. Future studies should address this question to identify the potential underlying biobehavioral mechanisms of the sleep-long COVID relationship.

In a recent study, Wang and collaborators^62^ showed that pre-infection psychological distress was prospectively associated with the incidence of various post-COVID-19 conditions. The similarity with our findings is unsurprising considering the close relationship between sleep disturbances and short sleep duration with psychological conditions like anxiety,^63,64^ depression,^63,65–67^ and stress disorders.^68,69^ Furthermore, a growing body of evidence supports a causal role of sleep problems in the occurrence of mental health problems,^65,70–72^ and treating sleep disturbances seems to improve the subsequent mental health outcomes.^73–75^ In this view, we could speculate that pre-infection sleep disturbances may get involved in the predictive role of psychological distress in the incidence of post-COVID-19 symptoms. In line with Wang and colleagues’ study,^62^ smell/taste dysfunction represented the only long-term symptom not prospectively associated with all previous sleep outcomes. However, as argued by those authors,^62^ the variability in anosmia incidence in long COVID may be explained by genetic or binding activity differences of the cell entry receptor for SARS-CoV-2 (the angiotensin-converting enzyme 2 receptor),^76^ and may be independent from inflammatory mechanisms potentially involved in the effects of distress and sleep. Further investigations should address this research question.

### Limitations and strengths

The present study has several strengths. The predictive effect of sleep variables was obtained by adjusting for important confounding factors (age, gender, and BMI) that were associated by previous studies with a higher risk for long COVID symptoms.^19–21^ Furthermore, the main findings were confirmed after controlling for the COVID-19 severity, which is an established predictor of long COVID conditions,^2,19,20,27^ but also a documented outcome of previous sleep problems.^77,78^ Third, the results were replicated in the same sample by using a different baseline assessment in December 2020. Finally, our findings were confirmed after controlling for the time elapsed from the baseline sleep assessment to the infection, suggesting a predictive role of sleep features even after several months. Nevertheless, it is worth noting that also the changes in sleep quality/quantity over time could affect the post-COVID-19 sequelae. In fact, variations in sleep variables between April and December 2020 were associated with the subsequent number of symptoms one month after infection (see Table S3 and Figure S3 in the supplemental material).

Several limitations should also be acknowledged. Notwithstanding our results rely on a longitudinal data collection, the long-term symptoms after COVID-19 were retrospectively reported by participants during the last follow-up assessment (April 2022). Second, the lack of evaluation of potential pre-existing health conditions and behaviors did not allow our analyses to account for other risk factors for post-COVID-19 conditions (e.g., type 2 diabetes, asthma, smoking status, physical activity). Moreover, sleep quality, insomnia symptoms, and sleep duration were evaluated using validated self-report instruments. Future longitudinal investigations should confirm our results using objective sleep-assessment instruments (actigraphy, polysomnography) and/or a clinical evaluation of sleep disturbances. Finally, our samples consisted of non-hospitalized adults and comprised a higher portion of female subjects, limiting the generalization of the results.

## Conclusions

In conclusion, this study suggests that pre-existing sleep disturbances and inadequate sleep duration are associated with subsequent risk of long-term symptoms after COVID-19. Our findings could have large-scale implications considering the sleep-loss epidemic in our society^79,80^ and the considerable rates of insomnia disorder and occasionally experienced insomnia symptoms among the adult population (10% and 20%, respectively).^81^ Furthermore, the present results may be even more relevant during a historical period that pervasively impacted the worldwide population’s sleep. Indeed, meta-analytic studies showed that half of the people experienced subthreshold and clinically significant insomnia symptoms in the first two pandemic years,^82^ and sleep disturbances affected four out of ten people worldwide.^83^ Raising public awareness about healthy sleep habits may represent an effective preventive approach to mitigate the COVID-19 repercussions in the long run, with substantial indirect effects at societal level. Further research is warranted to determine whether intervention aimed at promoting sleep quality/quantity could improve the long-term consequences of COVID-19.

## Data Availability

The data underlying this article will be shared on reasonable request to the corresponding author.

## Acknowledgments

We are grateful to all the Italians who participated in our study.

FS: Conceptualization, Methodology, Investigation, Data curation, Formal analysis, Writing - original draft, Writing - review & editing. GA: Investigation. DC: Investigation. LV: Investigation. ADA: Writing - review & editing. DT: Writing - review & editing. MF: Conceptualization, Methodology, Investigation, Supervision, Writing - review & editing.

## Data availability statement

The data underlying this article will be shared on reasonable request to the corresponding author.

