## Supplemental materials for "Poor sleep quality, insomnia, and short sleep duration before infection predict long-term symptoms after COVID-19"

Prof. Michele Ferrara, Ph.D.

Department of Biotechnological and Applied Clinical Sciences

University of L'Aquila

Via Vetoio (Coppito 2)

67100 Coppito (AQ)

Italy

### Supplemental Material

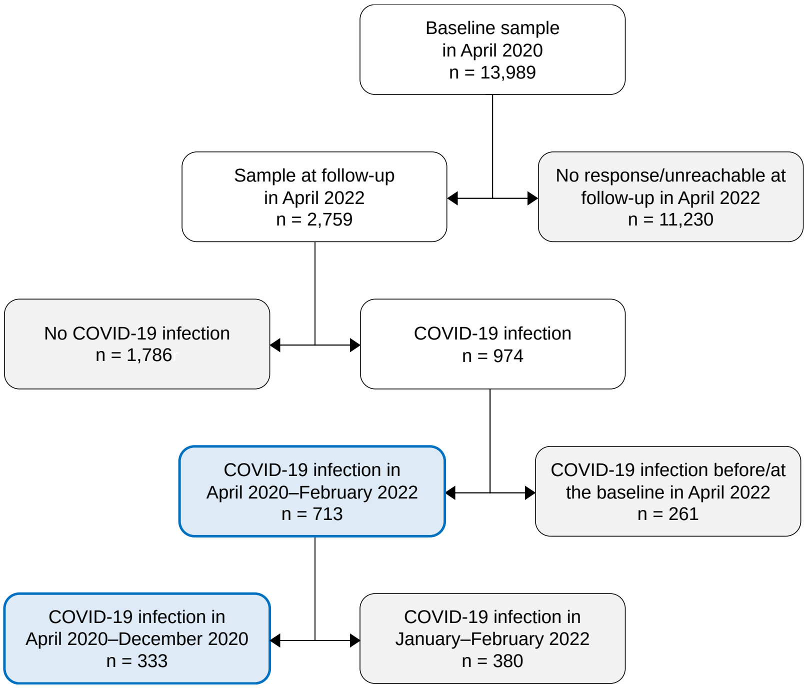


**Figure S1.** Flow chart of study participants.

*Notes:* Grey boxes indicated missing data or individuals excluded due to the study objectives. Blue box indicated analyzed samples. COVID-19 positivity information was collected at follow-up in April 2022.

**Table S1.** Information criteria (Akaike and Bayesian) for tested Negative Binomial and Zero-Inflated Negative Binomial models.

| Model | AIC | BIC |
|---|---|---|
| M1: sleep quality | | |
| NB | 3213.221 | 3240.229 |
| ZINB | 3174.956 | 3224.470 |
| M2: insomnia severity | | |
| NB | 3423.114 | 3450.531 |
| ZINB | 3390.338 | 3440.603 |
| M3: sleep duration | | |
| NB | 3452.988 | 3480.405 |
| ZINB | 3425.374 | 3475.638 |
| M4: sleep quality | | |
| NB | 1199.922 | 1222.399 |
| ZINB | 1182.043 | 1223.251 |
| M5: insomnia severity | | |
| NB | 1275.299 | 1298.147 |
| ZINB | 1250.948 | 1292.837 |
| M6: sleep duration | | |
| NB | 1285.192 | 1308.141 |
| ZINB | 1266.241 | 1308.131 |

*Notes:* A lower AIC or BIC value indicates a better fit.
*Abbreviations:* AIC, Akaike information criterion; BIC, Bayesian information criterion; M, Model; NB, Negative Binomial; ZINB, Zero-Inflated Negative Binomial.

*Supplemental statistical analysis*

First, we performed three supplementary zero-inflated negative binomial models (sM1, sM2, sM3) to estimate the effect of sleep variables (PSQI score, ISI score, TST, respectively) collected in December 2020 in predicting the number of long-term symptoms (range, 0–21) one month from COVID-19. These models were run on 209 individuals (mean age ± standard deviation, 35.34 ± 11.68 years; range, 18–82 years; 169 females) due to the smaller sample that participated in the last two assessments (December 2020 and April 2022), and because we only included people infected from December 2020 (after the questionnaire compilation) to February 2022. On the other hand, we cannot replicate predictive analyses on the number of symptoms three months after infection due to the inadequate sample size of participants (n = 32) infected from December 2020 to December 2022 that reported at least one long-term symptom.

Second, we evaluated if the occurred variations in sleep quality, insomnia severity, and sleep duration between April and December 2020 predicted the number of long-term symptoms by performing separate ZINB models that included baseline (April 2020) PSQI score (sM4), ISI score (sM5), or TST (sM6) and the respective change between the first two survey waves (e.g., Δ PSQI: PSQI in December 2020 − PSQI in April 2020) as independent variables. These models were performed only using the number of symptoms at one month from infection as dependent variable due to the inadequate sample size for studying long-term symptoms at three months from COVID-19 in the December 2020 sample.

Third, supplementary binomial logistic regressions were performed using the sleep variables collected in December 2020 as predictors of each symptom. Based on the event per predictor variable criterion adopted (at least five events), we analyzed only some symptoms that have been reported one month from infection (excluded symptoms: smell/taste dysfunctions, appetite reduction, depression, fever, PTSD, cardiovascular problems, OCD, and psychosis), while no three-month symptoms could be used as dependent variable due to insufficient events.

Fourth, supplementary binomial logistic regressions were performed to evaluate the association between the sleep characteristics collected in December 2020 and the recovery time to return to the pre-infection daily functioning level. However, only the odds of recovery time after 4 weeks could be tested due to insufficient number of individuals who reported recovering later than 12 weeks (n = 15). Therefore, the self-reported recovery time was entered as a dichotomous dependent variable (≤ 4 weeks vs. > 4 weeks) while sleep variables (PSQI score, ISI score, TST) were used as predictors. All the analyses described in this section were adjusted for age, gender, and body mass index (BMI; weight/height$^2$).

### *Supplemental results*

Analyses using sleep variables collected in December 2020 as predictors confirmed the effect of sleep quality, insomnia severity, and sleep duration on the number of symptoms one month from infection (Table S2). A one-unit increase in PSQI and ISI score, and a one-hour reduction of sleep duration predicted an increased number of symptoms by 11.4%, 6.2%, and 21.8%, respectively. The relationships between sleep variables and the number of long-term symptoms are depicted in Figure S2.

**Table S2.** Results from the negative binomial portion of the zero-inflated negative binomial regressions [*exp(B)*, 95% confidence intervals, *p*-value] estimating the effect of sleep variables (PSQI score, ISI score, TST) and confounding factors (age, gender, BMI) in December 2020 on the number of long-term symptoms one month after COVID-19.

| Predictor | sM1: sleep quality | | | sM2: insomnia severity | | | sM3: sleep duration | | |
|---|---|---|---|---|---|---|---|---|---|
| | *exp(B)* | 95% *CI* | *p* | *exp(B)* | 95% *CI* | *p* | *exp(B)* | 95% *CI* | *p* |
| Intercept | 2.209 | 0.981–4.974 | 0.056 | 2.785 | 1.180–6.574 | **0.019** | 34.383 | 9.470–124.838 | **< 0.001** |
| Gender* | 0.868 | 0.591–1.274 | 0.469 | 0.953 | 0.624–1.456 | 0.825 | 0.938 | 0.611–1.442 | 0.772 |
| Age | 0.983 | 0.971–0.995 | **0.006** | 0.986 | 0.974–0.999 | **0.035** | 0.981 | 0.968–0.995 | **0.006** |
| BMI | 1.023 | 0.991–1.055 | 0.162 | 1.021 | 0.988–1.056 | 0.216 | 1.014 | 0.980–1.050 | 0.420 |
| PSQI score | 1.114 | 1.076–1.153 | **< 0.001** | | | | | | |
| ISI score | | | | 1.062 | 1.035–1.089 | **< 0.001** | | | |
| TST (hour) | | | | | | | 0.782 | 0.699–0.875 | **< 0.001** |

*Notes*: *Female was used as reference for "*Gender*" factor; significant values are in bold.
*Abbreviations:* BMI, body mass index; CI, Confidence interval; ISI, Insomnia Severity Index; PSQI, Pittsburgh Sleep Quality Index; sM, Supplementary model; TST, Total sleep time.

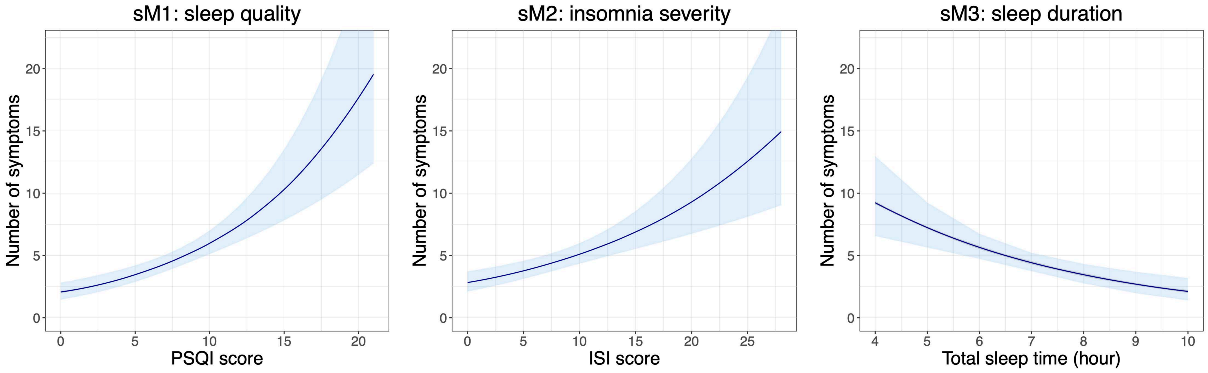


**Figure S2.** Relationships between sleep variables (PSQI score, ISI score, TST) in December 2020 and the number of long-term symptoms one month after COVID-19.
*Notes:* Light blue area represents 95% confidence intervals. Each model was adjusted for age, gender, and body mass index.
*Abbreviations:* ISI, Insomnia Severity Index; PSQI, Pittsburgh Sleep Quality Index; sM, Supplementary model; TST, Total sleep time.

Moreover, supplementary analyses (sM4–6) showed a significant association between the occurred variations in sleep variables from April 2020 to December 2020 and the number of symptoms reported one month after COVID-19 (Table S3). A one-unit increase in PSQI and ISI scores, and a one-hour reduction of TST in December 2020 compared to April 2020 predicted higher long-term symptoms after infection by 7.9%, 4.0%, and 15.1%, respectively. The relationship between Δ sleep variables and the number of symptoms after COVID-19 is shown in Figure S3.

**Table S3.** Results from the negative binomial portion of the zero-inflated negative binomial regressions [*exp(B)*, 95% confidence intervals, *p*-value] estimating the effect of sleep variable variations (Δ PSQI score, Δ ISI score, Δ TST) and confounding factors (age, gender, BMI, sleep variables collected in April 2020) on the number of long-term symptoms one month after COVID-19.

| Predictor | sM4: sleep quality variation | | | sM5: insomnia severity variation | | | sM6: sleep duration variation | | |
|---|---|---|---|---|---|---|---|---|---|
| | *exp(B)* | 95% *CI* | *p* | *exp(B)* | 95% *CI* | *p* | *exp(B)* | 95% *CI* | *p* |
| Intercept | 2.091 | 0.929–4.708 | 0.075 | 2.582 | 1.113–5.990 | **0.027** | 47.563 | 0.761–177.273 | **< 0.001** |
| Gender* | 0.925 | 0.631–1.356 | 0.690 | 0.944 | 0.624–1.427 | 0.784 | 0.954 | 0.627–1.450 | 0.825 |
| Age | 0.985 | 0.974–0.997 | **0.014** | 0.988 | 0.976–1.000 | 0.056 | 0.981 | 0.968–0.994 | **0.004** |
| BMI | 1.017 | 0.986–1.050 | 0.289 | 1.015 | 0.982–1.049 | 0.382 | 1.012 | 0.978–1.046 | 0.503 |
| PSQI score | 1.131 | 1.089–1.175 | **< 0.001** | | | | | | |
| Δ PSQI score | 1.079 | 1.037–1.124 | **< 0.001** | | | | | | |
| ISI score | | | | 1.082 | 1.050–1.114 | **< 0.001** | | | |
| Δ ISI score | | | | 1.040 | 1.009–1.072 | **0.012** | | | |
| TST (hour) | | | | | | | 0.995 | 0.993–0.997 | **< 0.001** |
| Δ TST (hour) | | | | | | | 0.849 | 0.742–0.971 | **0.017** |

*Notes*: Δ values are calculated subtracting values collected in December 2020 from those in April 2020. *Female was used as reference for "*Gender*" factor; significant values are in bold.
*Abbreviations:* BMI, body mass index; CI, Confidence interval; ISI, Insomnia Severity Index; PSQI, Pittsburgh Sleep Quality Index; sM, Supplementary model; TST, Total sleep time.

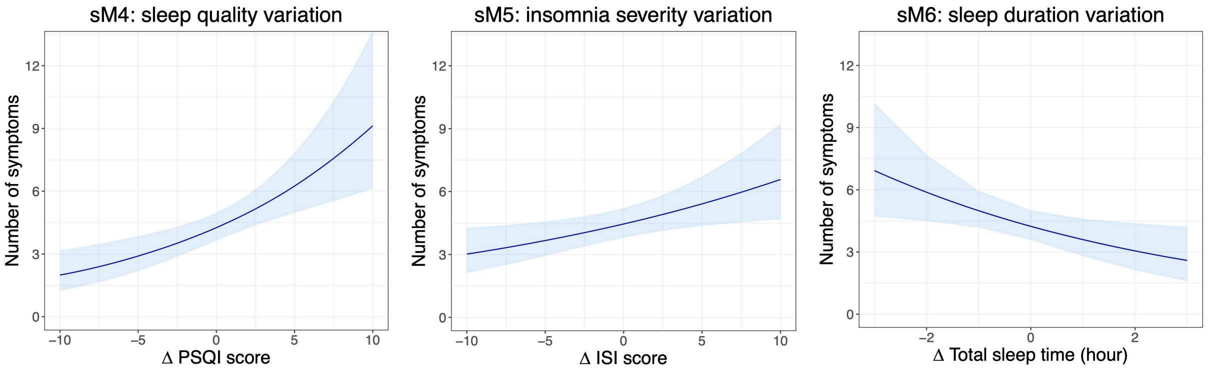


**Figure S3.** Relationships between variation in sleep variables from April 2020 to December 2020 and the number of long-term symptoms one month after COVID-19.

*Notes:* Δ values are calculated subtracting values collected in December 2020 from those in April 2020. Light blue area represents 95% confidence intervals. Each model was adjusted for age, gender, body mass index, and April 2020 scores.

*Abbreviations:* ISI, Insomnia Severity Index; PSQI, Pittsburgh Sleep Quality Index; sM, Supplementary model; TST, Total sleep time.

Supplementary binomial logistic regressions using the sleep variables collected in December 2020 as predictors of each symptom showed that lower sleep quality (Figure S4a) and more severe insomnia symptoms (Figure S4b) significantly predicted higher odds of all analyzed symptoms, except for persistent cough. Shorter sleep duration led to higher odds of over-tiredness, concentration/attention difficulty, breathlessness/dyspnea, headache, asthenia, sleep problems, anxiety, diffuse body pain, brain fog, and deterioration of perceived health status (Figure S4c).

Finally, logistic regressions using sleep variables collected in December 2020 as predictors confirmed the significant association of recovery in more than four weeks with PSQI [*OR* (95% *CI*) = 1.138 (1.043–1.241), $p = 0.004$] and ISI score [*OR* (95% *CI*) = 1.077 (1.018–1.138), $p = 0.009$]. No association was detected for sleep duration [*OR* (95% *CI*) = 0.854 (0.664–1.098), $p = 0.217$].

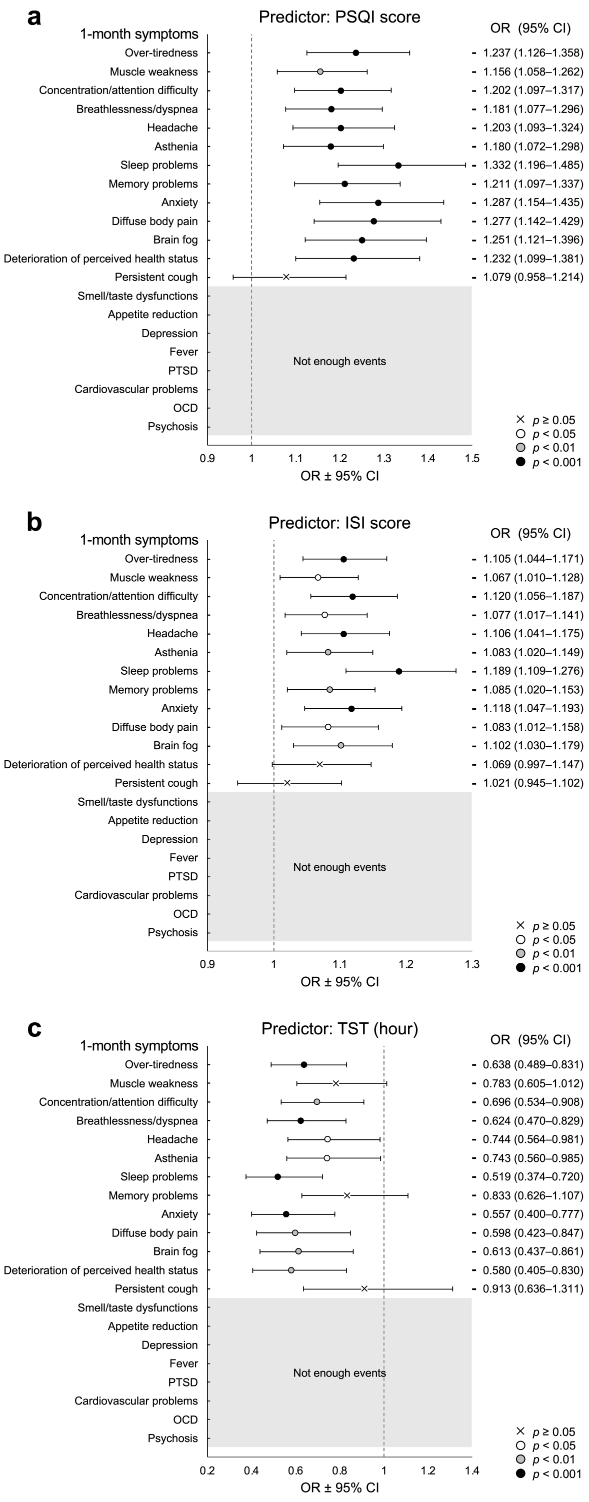


**Figure S4.** Results of logistic regressions (odd ratios and 95% confidence intervals) evaluating the predictive effect of sleep variables [PSQI score (a), ISI score (b), TST (c)] in December 2020 on the odds of each long-term symptom one month after COVID-19.

*Notes:* Long-term symptoms were ordered according to the prevalence data (top: most frequent) and represent the dependent variables. White dot indicates significance level at $p < 0.05$, grey dot at $p < 0.01$, and black dot at $p < 0.001$. "×" symbol indicates no statistically significant effect. Grey area indicates insufficient (< 5) outcome events per predictor. Each model was adjusted for age, gender, and body mass index.

*Abbreviations:* CI, Confidence Interval; ISI, Insomnia Severity Index; OCD, Obsessive-compulsive disorder; OR, Odd ratio; PSQI, Pittsburgh Sleep Quality Index; PTSD, Post-traumatic stress disorder; TST, Total sleep time.
